# Mask up! Testing strategies to increase mask usage in Kenya*

**DOI:** 10.1101/2022.02.16.22270815

**Authors:** Dennis Egger, Aleskandra Jakubowski, Carolyne Nekesa, Michael Walker

## Abstract

**Background:** COVID-19 continues to pose a major threat to countries around the world, and non-pharmaceutical interventions such as social distancing and face coverings remain important to reduce transmission, especially in settings with low vaccination rates. Despite a nationwide mask mandate in Kenya during the pandemic, proper masking remained low in Siaya County. We conducted a pilot study with the Siaya County Ministry of Health to improve mask adoption within Ugunja subcounty, and present initial findings on mask usage effects.

**Methods:** The study took place across 72 villages in Ugunja subcounty, which were randomly assigned to receive: (i) free mask and education on mask usage; (ii) only education on mask usage; or (iii) no mask or education by community health workers. A “role model” intervention was also cross-randomized across half of the villages, along with SMS messages reinforcing a variety of messages around masking. The intervention was administered in January 2021. Data collection was done via phone survey and direct observation of mask usage.

**Findings:** Preliminary analysis of the pilot study suggests providing free face masks may improve compliance, particularly in settings with higher COVID-19 risks. Two weeks to three months after the intervention, the free mask and education arm increased directly-observed correct mask usage by 3.1 percentage points (95% confidence interval 1.9, 6.0) on a control mean of 6.8 percent. Some treatment arms also improved COVID-19 knowledge and mask attitudes.

**Interpretation:** Interventions designed to increase adoption of health measures can be successful, but behavioral change is challenging and may require more intensive treatment.

## 1 Introduction

COVID-19 poses a major threat to countries around the world. Although some recommendations are conflicting, evidence suggests that face masks may significantly reduce the spread of SARS-CoV-2 and numerous governments, including Kenya, have issued mask mandates for certain scenarios. However, questions remain on how to effectively promote mask adoption: Despite masks being mandatory, recent evidence shows less than 20% wear them at markets in Western Kenya [3].

The Siaya County Ministry of Health (MoH) sought to improve mask usage amongst its citizenry in a cost-effective manner by considering a range of reasons why mask usage may be low: lack of access to affordable masks; misperceptions of the severity of COVID-19, its exponential growth, or of the effectiveness of masks; lack of salience or often forgetting masks at home; and/or social image concerns about wearing (or not wearing) a mask. We carried out a pilot randomized controlled trial (RCT) across 72 villages in Ugunja subcounty of Siaya of four different interventions designed to increase mask usage via different mechanisms. This involved: (i) provision of free masks and education about masks, (ii) mask education only, (iii) a role model intervention, and (iv) SMS messaging around masks. A main focus of this paper is on the effects of the masks and education arm on a) mask usage and b) mask attitudes.^1^

## 2 Methods

### 2.1 Intervention and Research Design

The study took place across 72 villages in Ugunja subcounty. Villages were randomly assigned to receive: (i) free mask and education on mask usage (24 villages); (ii) only education on mask usage (24 villages); or (iii) no mask or education (24 villages). In addition, half of the villages were assigned to a “role model” treatment, in which trusted community members served as advocates for mask usage. Some households also received additional messages: (a) details on mask effectiveness, (b) details on COVID-19 severity, and (c) reminders on bringing a mask. Appendix Figure A.1 outlines the full study design. The intervention was conducted in January 2021.

The masks for the intervention were provided by SafeHands Kenya, a private sector consortium deploying masks, soap and sanitizer across Kenya, and state #tibanisisi (We are the cure!) on the mask (see Figure A.2). In free mask and education villages, community health workers (CHWs) received training and guidance from the Ministry of Health on the educational scripts used for providing information to households. CHWs went house-to-house and provided education and either masks or soap in free mask vs education villages. Survey enumerators accompanied CHWs to record information on intervention implementation, and to provide the additional informational interventions. The role models in the role model treatment were identified pre-intervention by asking peers about individuals considered “trustworthy in health measures”. These individuals were invited to participate and incentivized to wear the #tibanisisi masks and encourage mask adoption in their social interactions and via text message.

### 2.2 Outcomes and Data Collection

Our main outcome measures are direct observations on a) whether a mask is visible and b) whether a mask is being worn properly (covering mouth and nose), as self-reports may overstate mask wearing [3]. Enumerators observed public spaces within villages from a safe distance and recorded the mask use, type and features of mask wearing by passer-by’s. We conducted multiple “waves” of observations, and within a wave villages were visited for three one-hour observation slots (morning, mid-day and evening). Pre-treatment (baseline) visits were conducted 4 months and 1 month prior to the intervention, 2 midline waves were conducted 1-4 weeks after the intervention, and 1 endline waves were conducted 5-8 weeks after the intervention. Thus, each village was visited 6 times prior to the intervention and 9 times after the intervention; a total of 69,503 direct observations were recorded within villages.^2^

We also look at whether the treatment changed household attitudes towards masks with data from phone surveys of households, specifically a) a COVID-19 knowledge index, comprised of questions about coronavirus spread, severity, and actions to reduce tranmission; and b) an index of attitudes about masks, namely their comfort level, social desirability and enforcement perceptions. Phone surveys took 15-30 minutes and were conducted concurrently with the direct observations (two rounds were collected pre-treatment and four rounds were collected post-treatment, roughly every two weeks after the start of the intervention).

IRB approval for the study was obtained from Kenya (Maseno University) and the US (University of California, Berkeley).

### 2.3 Statistical Analysis

We estimate the following linear regression for direct observation and phone survey data:

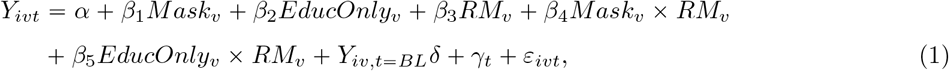

where *Y*_*ivt*_ is the outcome of interest for observation (respondent) *i* in village *v* at wave *t, Mask*_*v*_ is an indicator equal to 1 for free mask and education villages, *EducOnly*_*v*_ is an indicator for education only villages, and *RM*_*v*_ is an indicator for role model villages. For direct observations, the village (and corresponding treatment assignment) is defined based on the village in which observations are taking place (the observed individuals may or may not be from that village). The × operator indicates interaction terms, *Y*_*iv,t* = *BL*_ denotes baseline values of an outcome (included to improve statistical precision), and *γ*_*t*_ denotes survey wave fixed effects. We cluster standard errors at the village level.

We use the coefficients estimated from (1) to construct population-weighted average effects of (i) the free mask and education effect, (ii) the education only effect, and (iii) the role model effect, which we report below. We present results overall and over time.

## 3 Results

We begin by looking at trends in mask usage within our data, and treatment effects on mask usage as a result of the intervention, based on direct observations. We then turn to estimating the impact of the intervention on COVID-19 knowledge and attitudes using phone survey data.

### 3.1 Mask usage

We first look at effects on correct mask usage. Figure 1 uses the coefficient estimates from Equation (1) to calculate the mean free mask and education effect, education only effect, and role model effect, based on the share of individuals assigned to each in the population. We use pool data measured two weeks to three months after the intervention. The free mask and education arm increased mask usage by 3.1 percentage points (p-value = 0.037, 95% CI 1.9 pp - 6.0pp) from a mean correct mask usage rate in control villages of 6.8% during this period. Point estimates for the education only (1.5pp, 95%CI: -1.2pp-4.4pp) and role model (2.3pp, 95% CI: -0.5pp - 5.2p) interventions are positive, but not statistically significant at conventional levels.

**Figure 1:**
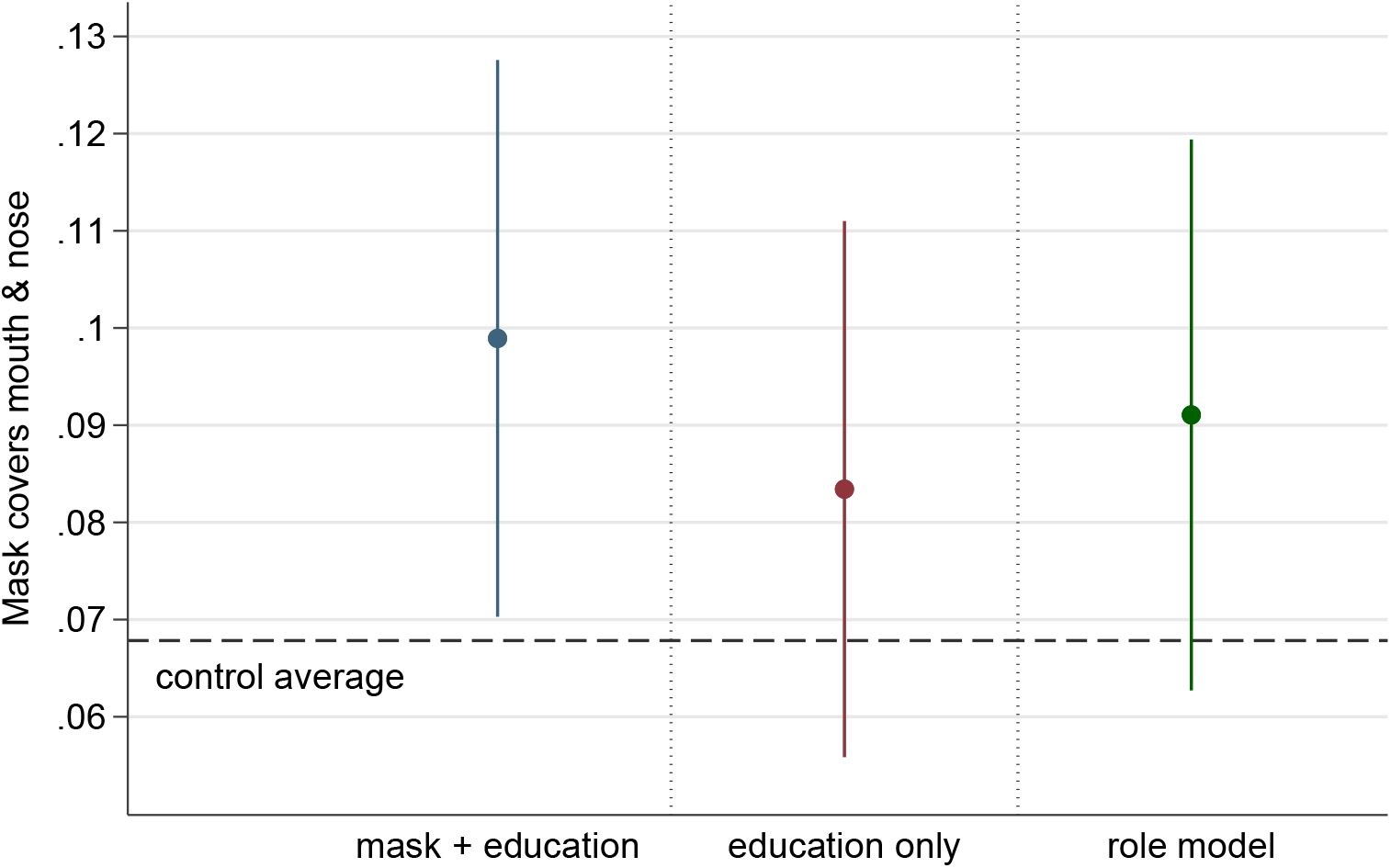
Intervention average treatment effects on correct mask usage *Source:* Direct observations data from two weeks to three months after intervention (February - April 2021). Intervention took place in January 2021.

There is some suggestive evidence that differences between treatment groups may diminish over time: at the last round of direct observations, conducted in April 2021, there is no longer a statistically significant difference between the free mask treatment group and the control group (Figure 2, Panel A), though point estimates are still positive (and the control group mean is higher compared to Figure 1). The smaller number of observations (as here we only use one round) may make this comparison underpowered. That said, positive effects are still observed in situations with the highest risk of transmission (indoors and on public transport, Figure 2, Panel B).

**Figure 2:**
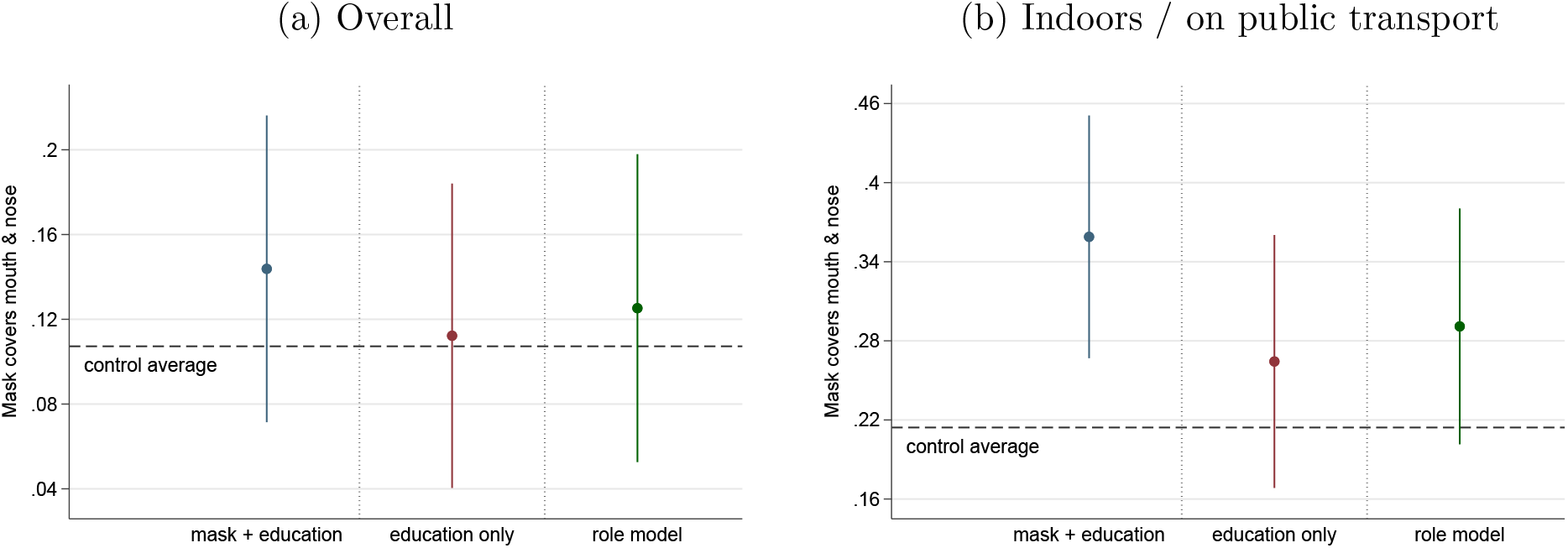
Intervention average treatment effects on correct mask usage, measured April 2021 *Source:* Direct observations data from April 2021. Intervention took place in January 2021.

**Figure 3:**
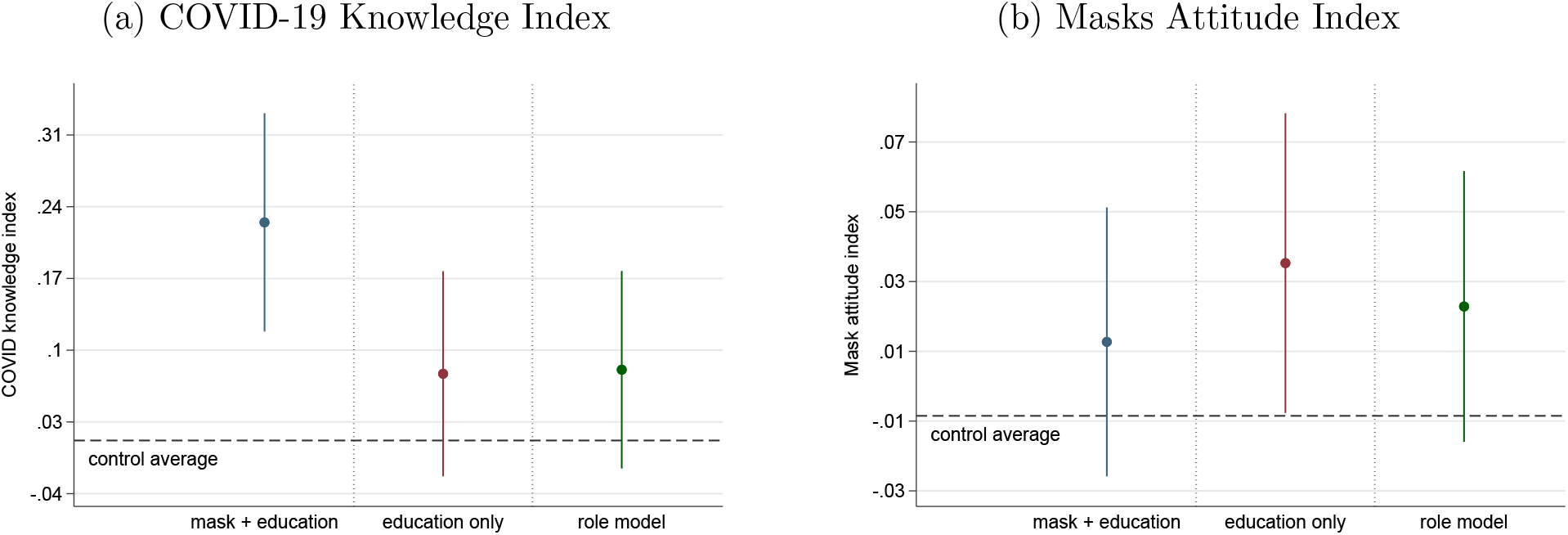
Intervention average treatment effects on COVID-19 knowledge and attitudes *Source:* Endline phone survey data. N=6,313.

### 3.2 Knowledge and Attitudes

Both the free masks and education only portions of the intervention included information about COVID-19. The free mask and education treatment resulted in a large increase in the COVID-19 knowledge index of 0.21 standard deviations (SE 0.054, p-value < 0.01). The education and role model treatment had positive estimated treatment effects (0.065 and 0.069, respectively), but these were not significant at traditional confidence levels. All treatment arms also resulted in positive point estimates for the masks attitude index, though the education only arm was the only arm with a statistically significant increase, providing some suggestive evidence that education can help change these attitudes. Note that these are “naive” p-values that do not take into account multiple hypothesis testing.

## 4 Discussion

These results suggest that the intervention was able to shift mask usage by a large amount in percent terms (nearly 50%), though the overall rates of proper mask usage still remained relatively low in this setting. We do see some suggestive evidence that education can lead to improvements in COVID-19 knowledge and attitudes around masks in certain arms. As education around masks could be provided by CHWs as part of their normal activities, this may be a cost-effective way to disseminate relevant information aroudn COVID-19 to households. While point estimates are generally positive for the role model intervention, it did not lead to statistically significant improvements in mask usage, knowledge or attitudes.

These gains are smaller in magnitude than other programs that have successfully increased mask adoption, such as [1] in Bangladesh. The intervention studied here was less intensive than the Bangladesh intervention, suggesting that more intensive interventions may be needed in order to change mask adoption behavior.

There are several limitations to this study. First, external validity: this study was conducted in one concentrated geographical area in Kenya during one phase of the pandemic. The villages in the study are rural, and thus more social interactions may take place outdoors, which could lower virus transmission risks. The study was also completed prior to the emergence of the omicron variant. The findings may not be applicable in other contexts or settings. Second, the two months of follow-up data provides a sense of the short-run effects of the program, but longer-term effects may vary. Third, the size of the study is smaller compared to other randomized trials that have studied mask adoption, such as in Bangladesh [1]. This study was designed to be more exploratory, testing a large number of intervention features that could potentially be scaled up, but this reduces statistical power for some of our comparisons.

## Data Availability

All data produced in the present study are available upon reasonable request to the authors

## Footnotes

1 Ongoing work uses the additional arms to test for mechanisms, particularly behavioral mechanisms, as pre-specified in our analysis plan (see A.3.

2 Direct observations were also collected from market centers, but we exclude these from this analysis; these will be used to understand spillover effects.

