## Supplement for "Mask up! Testing strategies to increase mask usage in Kenya*"

### A Supplemental Information

#### A.1 Supporting exhibits

Figure A.1: Experimental Design, village assignments

| **Total: 72** | Role model (36) | | | No role model (36) | | |
|---|---|---|---|---|---|---|
| No masks, no education (24)<br>*No households in these villages receive a visit, nor soap, nor mask, nor additional information* | 12 | | | 12 | | |
| | ***Additional Information Treatments***<br>*50% of households in these villages are randomly selected to receive additional information* | | | | | |
| | Mask effectiveness | Covid severity | Inattention | Mask effectiveness | Covid severity | Inattention |
| Education only (24)<br>*All households receive soap & in-person basic education* | 4 | 4 | 4 | 4 | 4 | 4 |
| Masks + Education villages (24)<br>*All households receive masks & in-person basic education* | 4 | 4 | 4 | 4 | 4 | 4 |

*Notes: Additional information treatments consist of an in-person visit and 4 weekly text message follow-ups. All 50% of households selected within a village receive the same additional information treatment.*

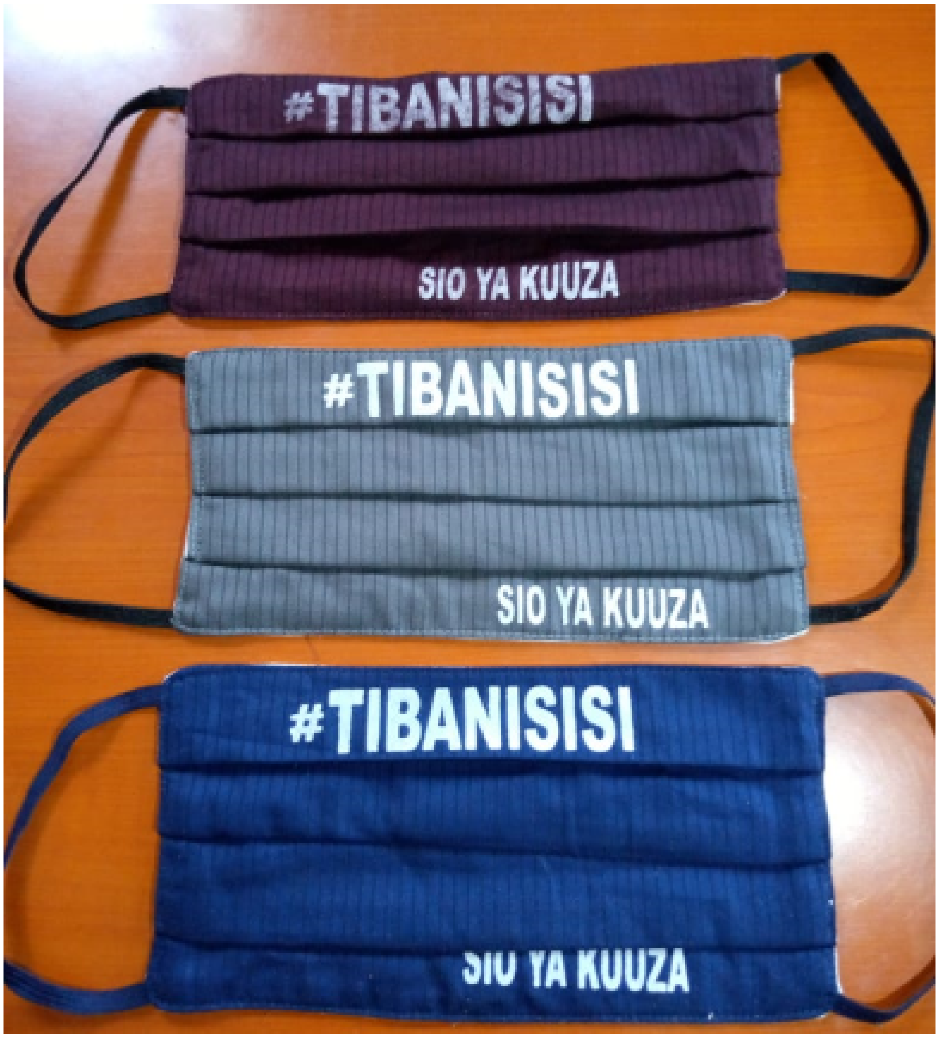


Figure A.2: Example of free masks provided by community health workers

*Notes:* #tibanisisi means "we are the cure" in Swahili; Sio ya kuuzu means not for sale.

#### A.2 Variable construction

Indicators for masks visible and masks used correctly are based on direct observation by survey enumerators for whether, for individuals that they publicly observed, masks (or another type of face covering) were a) visible, regardless of whether they were being worn properly, and b) whether masks were being worn over the nose and mouth.

The COVID-19 knowledge index is an inverse-covariance weighted index [2] of the following variables:

- Indicator for knowing how coronavirus spreads
- Indicator for believing coronavirus is more serious than malaria
- Indicator for correctly answering the age group at most risk of coronavirus
- Number of true / false questions about COVID-19 (generally misinformation circulating in the study area) answered correctly
- Indicator for providing key correct behaviors to reduce coronavirus transmission
- Indicator for not providing incorrect behaviors to reduce coronavirus transmission
- Indicator for washing mask among those who report having a mask

The index is normalized by the control group in each round.

The masks attitude index is an average of three subcomponents, where each subcomponent is an inverse-covariance weighted index (as above):

- Mask comfort
  - Indicator for disagreeing that masks are uncomfortable
  - Indicator for disagreeing that masks are unattractive
  - Indicator for not saying main reasons others do not wear masks is that masks are uncomfortable or don't look good
- Social desirability: the following questions are coded so that higher values indicate greater agreement with social desirability of mask usage:
  - I speak out when others around me do not wear masks
  - Others judge me for not wearing a mask in public spaces
  - God will judge those not wearing a mask
  - People that are not wearing masks are not good community members / citizens
  - People that do not wear face masks should pay a fine
- Enforcement perceptions: self-reported perceptions on being caught/reprimanded for not wearing a mask in the following locations, coded as 1=Never, 2=Rarely, 3=Sometimes, 4=Often, 5=Every time.
  - market center
  - religious gathering
  - on public transport
  - visit a store in your village
  - visit another household in your village

##### A.3 Pre-analysis plan

This study was pre-registered on the American Economic Association (AEA) Trial Registry: `https://www.socialscienceregistry.org/trials/6717`

The pre-analysis plan is available at the link, and includes additional details about the study and analyses that will be part of future work.
